# Immunogenicity of Pfizer mRNA COVID-19 vaccination followed by J&J adenovirus COVID-19 vaccination in two CLL patients

**DOI:** 10.1101/2021.09.02.21262146

**Authors:** Zoe L. Lyski, Sunny Kim, David Xthona Lee, David Sampson, Hans P. Raué, Vikram Raghunathan, Debbie Ryan, Amanda E. Brunton, Mark K. Slifka, William B. Messer, Stephen E. Spurgeon

## Abstract

**Importance:** Individuals with Chronic Lymphocytic Leukemia have significant immune disfunction, often further disrupted by treatment. While currently available COVID-19 vaccinations are highly effective in immunocompetent individuals, they are often poorly immunogenic in CLL patients. It is important to understand the role heterologous boost would have in patients who did not respond to the recommended two-dose mRNA vaccine series with a SARS-CoV-2 specific immune response

**Objective:** To characterize the immune response of two CLL patients who failed to seroconvert after initial mRNA vaccine series following a third, heterologous, COVID-19 vaccination with Ad26.COV2.S.

**Design:** Two subjects with CLL were enrolled in an IRB-approved observational longitudinal cohort study of the immune response to COVID-19 vaccination. After enrollment, they received a third vaccination with Ad26.COV2.S. Blood was drawn prior to original vaccination series, four weeks after mRNA vaccination, and again four weeks after third vaccination.

**Setting:** Eligible subjects were approached by oncologist overseeing CLL treatment and informed about study, at time of enrollment subjects consented to join the cohort study.

**Participants:** Sixteen subjects enrolled in the larger CLL cohort study, of whom two subjects received a third COVID-19 vaccination and were included in this analysis. Subject 1 is CLL treatment naive, while Subject 2 is currently on active treatment.

**Main Outcome(s) and Measure(s):** SARS-CoV-2 specific immune response, including plasma antibodies, memory B-cells, CD4 and CD8 T-cells were assessed prior to vaccination (baseline) as well as post vaccination series and post third dose.

**Results:** Of the two subjects who received Ad26.COV2.S doses, Subject 1 seroconverted, had RBD-specific memory B-cells as well as spike-specific CD4 T-cells while Subject 2 did not. Both subjects had a spike-specific CD8 T-cell response after original mRNA vaccination series that was further boosted after third dose (Subject 1), or remained stable (Subject 2).

**Conclusions and Relevance:** The results of this study, however small, is especially promising to CLL individuals who did not seroconvert following initial mRNA vaccination series. Especially those that are treatment naive, not currently in active treatment, or who may consider vaccination before beginning active treatment.

---

Chronic Lymphocytic Leukemia (CLL) is characterized by monoclonal proliferation of dysfunctional B-cells, leading to a broad range of immune defects. CLL patients face significant risk of morbidity and mortality from infections (1), including from SARS-CoV-2, the causative agent of COVID-19 (2). Vaccines can be instrumental in mitigating the risk of infections in CLL; however, responses to vaccination is highly variable and significantly influenced by CLL disease status, baseline characteristics, types of vaccine and active CLL therapy (3).

Although current COVID-19 vaccines elicit robust immunity in immunocompetent hosts (4), the antibody response in CLL patients is highly variable (5, 6) and particularly poor in patients with low total immunoglobulin levels, those that have had anti-CD20 monoclonal antibodies within the past year, or are undergoing active therapy with agents such as Bruton’s Tyrosine Kinase inhibitors (BTKi). The best responses to date have been in CLL patients who are in remission and/or years out from active treatment.

Given decreased vaccine efficacy in CLL, an additional dose of vaccine may be beneficial in CLL patients, especially given rise of variants of concern (VoCs). Initial data from solid organ transplant recipients on immunosuppression showed a role for additional vaccination (7), leading the FDA to extend the EUA for Pfizer-BioNTech and Moderna mRNA vaccines to include additional doses in immunocompromised patients. However, the results in solid organ transplant patients may not be generalizable to CLL, and additional studies are needed to better define vaccine responses in the CLL patient population, including the role of mixing mRNA vaccination with other vaccine formulations, such as the adenovirus vectored vaccine Ad26COV2.s, commonly known as Johnson and Johnson (J&J) vaccine.

Here we describe two CLL patients who “self-referred” to outside pharmacies for an additional vaccination with J&J COVID-19 vaccine following 2 doses of the BNT162b2 vaccine (Pfizer-BioNTech). Both patients had previously enrolled as study subjects in an IRB approved observational study, (OHSU IRB# 21230) to investigate immune response following COVID-19 vaccination. The additional J&J dose was subsequently self-reported to the study team. On initial enrollment, demographics, CLL disease characteristics, and treatment details were collected (Table 1), and baseline laboratory values were obtained, included semi-quantitative SARS-CoV-2 spike antibody titer, serum IgG, a complete blood count, and multicolor flow cytometry measuring immune cell populations (Table 1). Whole blood was collected for additional serologic and cellular studies.

**Table 1.** Baseline Characteristics and demographics for subjects included in the study. Normal ranges for each of the B-cell subset are in parenthesis under each B-cell type.

| Subject ID | Age | Gender | Year of Diagnosis | Current Treatment | Prior Treatment | IgG mg/dL | Absolute Lymphocyte Count K/mm3 | B-cells (CD19+) % | Naïve B-cells (IgD+CD27-) % | Memory B-cells (IgD-CD27+) % | B1 B-cells (CD5+CD19+) % |
| --- | --- | --- | --- | --- | --- | --- | --- | --- | --- | --- | --- |
|  |  |  |  |  |  | (768 - 1632) | (1.00 - 4.80) | (4-17) | (50-80) | (5-21) | (<6) |
| 1 | 60's | F | 2014 | None | None | 834 | 21.09 | 76 | 0.092 | 59.1 | 76.18 |
| 2 | 80's | F | 2014 | Ibrutinib | Obinutuzumab | 510 | 5.93 | 61 | 11.37 | 0.45 | 59.96 |

SARS-CoV-2 spike receptor binding domain (RBD)-specific antibody levels were tested by ELISA and endpoint titers were calculated as previously described (8). In addition, baseline PBMC samples were functionally tested for the presence of SARS-CoV-2 spike RBD-specific memory B-cells (MBCs) by limiting dilution assay (9, 10) and CD4+ and CD8+ T-cells were functionally assessed for the presence of IFN*γ* and TNF*α* secretion following spike protein derived peptide stimulation.

Neither subject had pre-vaccination B-cell responses as measured by RBD-specific antibodies or MBCs. Neither had a virus-specific CD8+ response at baseline. While Subject 2 had spike peptide-reactive CD4+ T-cells at baseline these cells were unresponsive and did not expand following vaccination. In contrast, CD8+ responses were observed after mRNA vaccination in both subjects (Fig. 1). It has previously been reported that SARS-CoV-2 naïve individuals may have preexisting cross-reactivity to SARS-COV-2 peptides through prior infection by common cold coronaviruses: SARS-CoV-2 specific CD4+ T-cells have been identified in 20-50% of people without SARS-CoV-2 exposure or vaccination (11).

**Figure 1.**
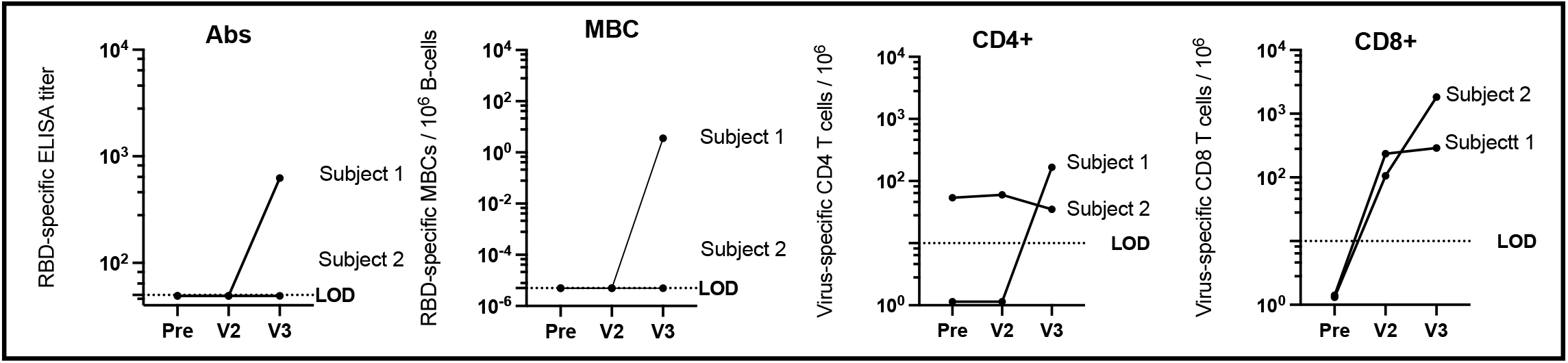
Immune response to COVID-19 vaccination in CLL subjects. RBD-specific antibody (Ab) titer. Subjects without a detectable Ab titer (< 1:50 serum dilution) were assigned a value of 49. Frequency of RBD-specific MBCs per 10^6^ CD19+ B-cells following ex vivo stimulation. Subjects who did not have a detectable response were assigned a value of 5×10^−6^. SARS-CoV-2 spike peptide-reactive CD4 and CD8 T-cells are defined as double positive for IFN*γ* and TNF*α* cytokine secretion. Patients who did not have a detectable T-cell response were assigned an arbitrary number between less than 2. Visit 1 (pre) blood draw was taken 21 and 40 days prior Pfizer vaccine series (2-doses). Visit 2 (V2) blood draw was taken 33 and 24 days post vaccination, and visit 3 (V3) was drawn 30 and 27 days after 3rd vaccination with J&J.

Approximately four weeks after initial vaccination neither subject had detectable RBD-specific SARS-CoV-2 antibodies or MBCs. Both had measurable vaccine-induced CD8+ T-cell responses following mRNA vaccination, although CD4+ responses did not appear to increase above baseline (Fig. 1).

Subject 1 received the J&J vaccine 104 days and Subject 2-81 days after completion of the BNT162b2 vaccine series. Following J&J vaccination additional samples were obtained, Subject 1, 30 days after third vaccine, and Subject 2, 27 days following third vaccine. Interestingly, Subject 1 had undetectable RBD-specific antibodies, RBD-specific MBCs, and virus-specific CD4+ T-cells after initial vaccination series. However, following an additional vaccination, all three measures increased above the limit of detection, RBD ELISA titer of 625, RBD-specific MBC frequency of 3.6 / 10^6^ B-cells, and 166 spike-specific CD4+ T-cells / 10^6^, and a spike-specific CD8+ T-cell response that remained stable and did not boost appreciably following 3^rd^ vaccination (Fig. 1). Subject 2 did not seroconvert or have detectable virus specific MBCs after their primary mRNA vaccination series however they had a spike-specific CD8+ T-cell response that further boosted after a 3^rd^ dose and a virus-specific CD4+ response that didn’t change following original vaccine series or 3^rd^ dose of J&J.

Other than subject age (60s vs 80s), the most notable difference between the subjects’ baseline characteristics (table 1) is that Subject 1 was treatment naive, while Subject 2 had undergone previous treatment (6 years ago) with obinutuzumab an anti-CD20 mAb and is currently on active treatment with Ibrutinib, since 2017. Both had baseline B-cell frequencies outside of the normal range, with Subject 1 exhibiting a low percentage of naïve B-cells (0.092) and a high percentage of MBCs (59.1), while Subject 2 had a low percentage of naïve B-cells (11.37) and MBCs (0.45). Although Subject 2 had mild hypogammaglobulinemia, neither had a history of recurring infections or need for IgG supplementation. Levels of baseline CD4+ and CD8+ T-cells (absolute values) were also normal, in each subject prior to vaccination (data not shown). Both had very low percentages of naïve B cells which could explain the initial poor response to vaccination. The significance of the increased percentage of MBCs in Subject 1 is unclear but does suggest some broader preservation of normal B cell maturation and immune function.

Although Subject 1 did have an immune response, antibody levels were relatively low as compared to some of the levels observed in immunocompetent post-vaccine populations (12) and certain CLL populations (5). The clinical significance of specific antibody levels remains unknown.

Active treatment with Bruton’s Tyrosine Kinase (BTK) inhibitors like ibrutinib may have a profound impact on B-cell survival, differentiation, and production of antibodies as the absence of intact BTK–dependent B-cell receptor mediated signaling prevents B-cells from differentiating into mature peripheral B-cells. Immune response following vaccination or natural infection is limited in these patients (13). Recall to antigens encountered prior to treatment appears to remain largely intact, however response to novel antigens encountered during treatment seems to be abrogated. Subject 2 has been on ibrutinib for over four years. The impact of prolonged treatment vs. shorter-term BTK inhibition on immune responses is unknown.

However, clinical data (14) suggest some improvement in humoral immunity with prolonged (> 6 months) treatment. T-cells are also disrupted in individuals with CLL and even further disrupted with BTK treatment (15). In the cases presented here both subjects did have an increase in virus specific CD8+ T-cells however the significance is unclear in terms of protection as neutralizing antibodies are often viewed as the correlate of protection against COVID-19. The results of this study, however small, provide initial evidence that a 3^rd^ vaccination against COVID-19 with the heterotypic vaccine Ad26COV2.s results in an immune response that was not observed following the recommended 2-dose mRNA vaccination series. This is especially promising news to subjects who are treatment naïve, not currently in active treatment, or who may consider vaccination before beginning active treatment.

## Data Availability

No additional data or external datasets.

## Acknowledgements

The authors would like to thank the subjects for participating in this research study.

